# Active Bilingual Immersion Leads to Active Brain Cleansing: Multimodal Evidence for L2 Engagement Optimizing Glymphatic Function

**DOI:** 10.64898/2026.03.18.26348672

**Authors:** Ruisi Wang, Qiwei Guo, Xinglin Zeng, Chantat Leong, Cheneryue Zhang, Yupeng Zhang, Jubin Abutalebi, Andriy Myachykov

## Abstract

**Background:** The brain’s glymphatic system plays a vital role in maintaining neural health. However, little is known about whether second language (L2) immersion can influence this clearance pathway.

**Methods:** 50 high-proficiency L2 English speakers (mean age: 32.6 years; 78% female) were assessed for glymphatic function using three multimodal MRI markers: BOLD-CSF coupling strength (fMRI), choroid plexus ratio (structural MRI), and DTI-ALPS index (diffusion MRI). Analyses examined relationships between glymphatic markers and L2 immersion duration, age of acquisition (AOA), and active use environment, controlling for age, education, and sex.

**Results:** L2 immersion duration correlated significantly with better glymphatic function. Longer immersion related to better BOLD-CSF coupling strength (r = −0.315, p < 0.05) and decreased choroid plexus ratios (r = −0.39, p < 0.05), suggesting enhanced brain-CSF coordination and fewer pathological CSF production structures. Mediation analyses demonstrated that immersion influenced ALPS indirectly through effects on choroid plexus morphology and BOLD-CSF coupling. L2 AOA moderated the immersion-coupling relationship: individuals who began learning after age 9.53 showed stronger associations between immersion and BOLD-CSF coupling, though AOA did not moderate choroid plexus effects. As for L2 immersive active is associated with better glymphatic function, while L2 immersive passive and L2 non-immersive active are both unrelated.

**Conclusions:** L2 immersion associates with better glymphatic system function through multiple pathways, including improved brain-CSF coordination, optimized choroid plexus structure, and increased perivascular flow. These findings provide novel neurobiological evidence that bilingual experience may confer neuroprotective benefits through brain waste clearance mechanisms.

## Introduction

While the impact of bilingualism on brain structure and function is well-documented, it is increasingly recognized as a form of sustained cognitive exercise that actively shapes brain health. In parallel, the glymphatic system—a waste clearance pathway facilitating cerebrospinal fluid (CSF) and interstitial fluid (ISF) circulation^1,2^ —has emerged as a vital contributor to brain maintenance. Beyond its role in clearing neurotoxic waste to prevent neurodegeneration^3–9^,, glymphatic function is now understood to play a critical role in high-level cognitive processes, including learning and memory^10–14^. Despite the established influence of both bilingual engagement and glymphatic integrity on brain function, the relationship between bilingual experience and glymphatic function remains entirely unknown. Investigating this intersection is essential to determining whether the glymphatic system serves as a novel physiological substrate for the cognitive reserve associated with bilingualism.

### Glymphatic system and brain health

The glymphatic system operates as an integrated CSF pathway comprising three interconnected components^15^: CSF production^16^, circulation dynamics^17,18^, and waste clearance through perivascular channels^19,20^. The choroid plexus serves as the primary CSF production site^16^, residing within the brain’s ventricular system – specifically the lateral, third, and fourth ventricles – and consisting of epithelial cells enveloping dense vascular networks that continuously generate CSF to initiate the glymphatic circulation cycle^16,21,22^. CSF circulation operates via neurovascular coupling, where neural slow waves induce cerebral blood volume fluctuations that generate compensatory CSF flow under constant intracranial volume constraints^23,24^. The BOLD-CSF coupling index quantifies this process by measuring synchronization between blood-oxygen-level-dependent (BOLD) signals and CSF flow patterns, providing a window into circulation efficiency^17,18^. The final component involves waste removal through perivascular spaces (PVS)^25^ – fluid-filled channels surrounding blood vessels that facilitate metabolite clearance from brain tissue. The diffusion tensor imaging-based Analysis along the Perivascular Space (ALPS) index offers an indirect assessment of glymphatic clearance capacity by measuring water molecule diffusion patterns along perivascular space orientations, thereby indicating how effectively the system removes waste products^19,20^. Together, these three components – production, circulation, and clearance – form a complete functional network that maintains brain health through continuous waste removal and fluid exchange.

In recent studies, the directional pathological changes of these three indicators in neurodegenerative diseases show remarkable consistency. Specifically, neurodegenerative conditions manifest as choroid plexus enlargement^26^, decreased BOLD-CSF coupling intensity^17,27^ indicating reduced synchronization between brain activity and CSF flow, and decreased ALPS index values^28^ reflecting diminished perivascular space flow strength.

Choroid plexus enlargement may impair CSF clearance function and represent an inflammatory response associated with disease progression. These convergent patterns across multiple glymphatic markers suggest that disrupted waste clearance mechanisms play a fundamental role in neurodegenerative pathology, with compromised CSF production structures, impaired circulation dynamics, and reduced perivascular clearance capacity collectively contributing to the accumulation of toxic metabolites that characterize conditions such as Alzheimer’s disease and other forms of neurodegeneration. The consistency of these directional changes across different measurement approaches strengthens the evidence that glymphatic dysfunction represents a core pathophysiological feature rather than a secondary consequence of neurodegeneration^10^.

In the context of normal aging, Choroid plexus volume demonstrates significant annual increases (14.92 mm³/year)^11^,suggesting progressive structural changes over time. Furthermore, larger ChP volume is associated with poorer performance across multiple cognitive domains, including scores on the Mini Mental State Examination (MMSE)^11^, revised Addenbrooke’s Cognitive Examination (ACE-R)^12^, and verbal fluency tasks^10^. While these findings primarily emerge from studies of aging and pathological populations, the relationship between choroid plexus volume and experience related activities such foreign language experience in healthy young adults remains unknown. Also, weakened BOLD-CSF coupling strength (reflected by increased coupling values) indicates disrupted coordination between CSF flow and neural activity, indicating impaired clearance of amyloid-beta (Aβ) and tau proteins that accumulate in neurodegenerative diseases^17^. Decreased ALPS index values signal reduced water diffusion along perivascular pathways, suggesting compromised waste removal capacity through these critical clearance channels^29^. The convergent directionality of these three markers – enlarged choroid plexus structures, weakened brain-CSF synchronization, and diminished perivascular flow – points to a unified pattern of glymphatic dysfunction underlying neurodegeneration. This consistent profile across multiple independent measures strengthens the interpretation that impaired waste clearance represents a core pathophysiological mechanism rather than an epiphenomenon of disease. However, most research has focused on clinical and aging populations, leaving a significant gap in understanding whether cognitive experiences such as second language immersion might influence these glymphatic parameters in younger, neurologically healthy individuals who have not yet developed pathological changes^17^ and accelerated cognitive decline^13^.

Furthermore, all three glymphatic biomarkers mentioned above show potential relationships with language function. In patients with mild cognitive impairment, verbal memory impairment is an early core symptom, and the larger the right choroid plexus volume, the worse the verbal learning memory ability^30^. And primary progressive aphasia shows reduced BOLD-CSF coupling intensity versus healthy controls^27^. In addition, children with language developmental delays show decreased ALPS indices^31^, and the ALPS index is modulated by language lateralization^32^. At the same time, research on its relationship with multilingual experience is lacking. In summary, while all three biomarkers are validated, each has limitations: ChP only assesses choroid plexus tissue, BOLD-CSF provides vascular ^33,34^ dynamics but lacks morphological information, and ALPS is limited to lateral ventricle level without reflecting whole-brain function. Given the lack of a gold standard for glymphatic function^35^, multimodal approaches warrant exploration to capture the relationship with bilingual experience.

### Bilingualism experience and brain plasticity

Evidence for bilingual experience affecting brain plasticity has been well documented across multiple neuroimaging studies^36^. Brain plasticity refers to the brain’s capacity to reorganize itself^37^ in response to environmental changes^38^. The use of language as the primary medium for human interaction with the external environment^39^ leads to the changes in the brain structure, which are both long-lasting^40–43^ and extensive in scope^44,45^. Bilingual individuals demonstrate structural and functional brain changes in regions associated with language processing^46–48^, executive control^49–53^, and cognitive flexibility^54,55^, including increased gray matter density in the anterior cingulate cortex^56,57^, inferior parietal lobule^58,59^, and prefrontal regions^60^. These neuroplastic adaptations extend beyond language-specific areas to encompass broader neural networks involved in attention, inhibitory control, and task switching^46,61^. Connectivity studies reveal that bilinguals exhibit enhanced network efficiency and reorganized connectivity patterns^62–66^, suggesting that sustained language switching and management demands drive widespread neural remodeling. Furthermore, longitudinal studies of second language learning demonstrate that even relatively brief immersion experiences can induce measurable structural changes^67–70^, with effects varying according to proficiency level, age of acquisition (AOA), and intensity of language use. Given that neuroplasticity mechanisms can influence various aspects of brain physiology^71^, including vascular health, inflammatory responses, and metabolic efficiency, it remains plausible that bilingual experience might extend its effects to the glymphatic system, potentially modulating CSF production, circulation dynamics, and waste clearance pathways through these broader neurophysiological adaptations.

Learning a foreign language to fluency^72,73^ or relocating to a second language environment^34,67^ represents enormous cognitive challenges, and such substantial environmental stimulation reshapes brain systems through sustained neuroplastic adaptation. With the development of neuroimaging technologies, research on bilingual experience and brain plasticity shows two notable trends.

First, research has evolved from binary monolingual-bilingual comparisons to examining continuous language experience variables (AOA^74^, immersion duration^75–78^, proficiency^74^, usage patterns and so on^79–81^) that better capture the multidimensional nature of bilingual experience and allow identification of dose-response relationships with neural outcomes^80,82,83^. Early studies found bilinguals showed greater gray matter^84,8523^, higher white matter integrity^86,87^, and stronger functional activation^33,42^ in executive function^422288^ and language control networks compared to monolinguals. However, inconsistent findings in subcortical neuroplasticity^79^ have led recent studies to increasingly treat language experience as continuous rather than categorical variables.

Second, the definition of brain plasticity has evolved from early Voxel-Based Morphometry (VBM) volume studies to broader neuroplasticity indicators, as a meta-analysis of eight VBM studies found no consistent structural differences between bilinguals and monolinguals^89^. This inconsistency likely stems from the non-linear nature of gray matter changes, which can increase or decrease depending on developmental stage, learning phase, and neural processing efficiency, making volumetric measures problematic for cross-sectional brain reserve assessment^80,81,90,91^. Researchers now employ diverse methodological approaches including functional network connectivity^64,65^, basal ganglia metabolic concentrations^92^, white matter microstructural integrity through diffusion imaging^86,87^, and neurotransmitter system functioning via magnetic resonance spectroscopy^64,93^ to identify brain benefits of bilingual experience.

In summary, following current research trends by employing continuous language experience variables and novel neuroimaging methods to assess brain plasticity enables deeper understanding of how foreign language learning and immersion experiences impact brain health across multiple physiological systems.

### The role of immersive L2 environment

Second language (L2) immersion is a basic L2 variable and global issue. Over 200 million international migrants worldwide face immersive foreign language environments, while short-term experiences like studying abroad are also increasingly common^94^. L2 immersion environments provide qualitatively different neural stimulation compared to classroom-based or non-immersive learning contexts^95,96^. In L2 immersive environments, learners receive continuous, authentic auditory and visual linguistic stimuli while facing pressure for active L2 use. As a result, the brain undergoes adaptive changes in response to these environmental demands with L2 immersion not only enhancing language proficiency and executive functions in young popluations^95^, but also offering protection against age-related language decline in elderly individuals^43^.

In neuroimaging studies, immersive bilingual environments create distinct patterns of brain plasticity^67,97,98^ across subcortical structures, cortical regions, and white matter tracts, with immersion duration proving to be more critical than proficiency levels^34,67^. Immersion experiences, characterized by daily necessity to navigate social, professional, and practical situations in the second language, impose sustained cognitive demands that extend beyond classroom-based instruction or limited conversational practice^96, 99,100^. These demands include real-time comprehension under varied acoustic conditions^100^, rapid lexical access without translation mediation^101^, pragmatic adjustment to cultural communication norms^102^, and continuous monitoring of linguistic context to select appropriate language modes^103^.

Neuroimaging evidence suggests that immersive environments produce more pronounced structural and functional adaptations than equivalent hours of formal instruction, with effects observed in executive control networks, language processing efficiency, and cross-linguistic interference management^104–106^. Studies comparing immigrants who use their second language daily with classroom learners of similar proficiency reveal differential patterns of white matter connectivity^104,105^, and functional activation pattern^107^, suggesting that the ecological validity and communicative pressure inherent to immersion environments drive neuroplastic changes through mechanisms distinct from explicit learning. Furthermore, the duration and intensity of immersion experience show dose-dependent relationships with neural outcomes^105^, indicating that prolonged exposure to linguistically demanding environments produces cumulative adaptations that may extend to broader cognitive and physiological systems beyond language processing itself.

Although L2 immersion duration affects brain plasticity and serves as a brain health protective factor, it essentially reflects mere temporal exposure while overlooking actual engagement levels^98^. Some individuals may maintain native language habits even within immersive environments through ethnic enclaves, native language media consumption, or social networks that minimize L2 demands^108^, while others actively adapt by seeking out L2 interactions, engaging with local communities, and deliberately practicing the target language across diverse contexts^109,110^. Whether L2 immersion itself or the active use within it drives neural effects remains unclear, as physical presence in an immersive environment does not guarantee meaningful linguistic engagement. Since L2 active use is also a duration-based variable measured in time spent communicating^98^, understanding its specific positive contributions beyond passive environmental exposure requires further investigation to disentangle the effects of opportunity from actual engagement. Additionally, our understanding of how AOA and immersion duration interact to reshape the brain remains limited. The critical period hypothesis suggests that early language learning occurs through different neural mechanisms than adult acquisition^111–113^, yet whether late bilinguals require greater neural compensation or demonstrate different plasticity patterns than early bilinguals when facing equivalent immersive challenges remains unclear^113–117^. The AOA effect suggests that the timing of language onset serves as an “organizing variable” that triggers environmental and cognitive shifts^118^. Consequently, late learners may recruit additional cognitive resources to overcome L1 entrenchment or employ distinct processing strategies to achieve comparable functional outcomes. This may lead to differential effects of immersion duration based on the age of onset, though empirical evidence characterizing these interactions remains emergent.

Based on the evidence presented, investigating the relationship between bilingual experience and glymphatic function addresses a critical gap at the intersection of language neuroscience and brain health mechanisms. While bilingualism’s neuroprotective effects against cognitive decline and neurodegeneration are increasingly recognized^119–125^, the physiological pathways underlying these benefits remain poorly understood. The glymphatic system represents a promising candidate mechanism, given its central role in waste clearance and its documented dysfunction in neurodegenerative conditions that bilingualism appears to protect against.

Moreover, existing bilingualism research has largely focused on neural structural and functional changes, overlooking non-neuronal systems that fundamentally support brain health. By examining whether L2 immersion experience influences choroid plexus morphology, brain-CSF coupling dynamics, and perivascular flow efficiency, this study extends bilingualism research beyond traditional cognitive and structural neuroimaging to encompass fundamental physiological processes. Furthermore, clarifying how AOA moderates these relationships and distinguishing active engagement from passive environmental exposure addresses theoretical questions about critical periods and ecological validity that have broader implications for understanding experience-dependent brain adaptation. Given that hundreds of millions of people worldwide navigate immersive bilingual environments, establishing whether such experiences confer measurable benefits to brain waste clearance systems would provide novel insights into accessible, non-pharmacological approaches to promoting brain health across the lifespan.

### Study Design

As shown in Figure1, our study objectives are threefold: first, to establish the connection between bilingual experience and glymphatic system function as a novel neuroprotective pathway; second, to examine how age of acquisition moderates this relationship in accordance with in accordance with established AOA effect frameworks^118^; and third, to clarify the specificity of active versus passive L2 use within immersive environments and their differential effects on brain health mechanisms.

#### Hypothesis 1: L2 Immersion Enhances Glymphatic Function

We hypothesize that L2 immersion duration will be positively associated with glymphatic function, evidenced by enhanced BOLD-CSF coupling strength, optimized choroid plexus morphology (smaller volume ratios), and increased perivascular flow dynamics (higher ALPS index values). This hypothesis posits that sustained cognitive demands and neural adaptations resulting from immersion experiences promote more efficient waste clearance mechanisms across all three glymphatic components.

#### Hypothesis 2: Moderation by L2 AOA

We hypothesize that L2 age of acquisition will moderate the relationship between immersion duration and glymphatic function, with association patterns varying depending on when language learning began. This hypothesis is grounded in age effect frameworks suggesting that the timing of acquisition influences the degree of L1 entrenchment^118^, potentially leading to differential neuroplastic responses to immersion experiences., potentially leading to differential responses to immersion experiences in terms of physiological adaptation.

#### Hypothesis 3: Active Immersion Specificity Hypothesis

We hypothesize that only active L2 use within immersive environments, rather than passive immersion exposure or active L2 use in non-immersive contexts, will significantly associate with enhanced glymphatic function. This reflects the synergistic effects of meaningful engagement combined with ecological linguistic demands.

**Figure 1.**
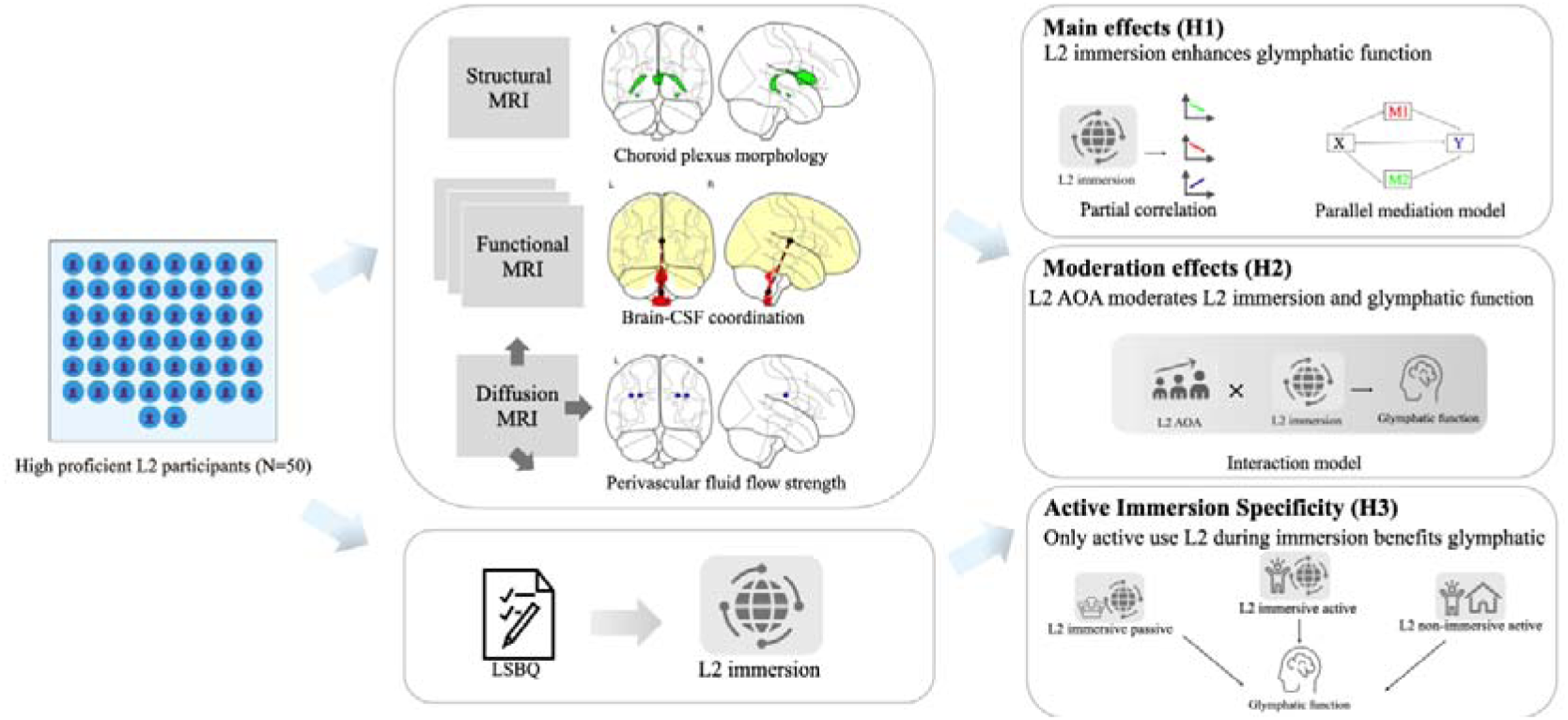
Research procedure

## Methods

### Data Source, Participants, and Ethics

All procedures performed in this study were in accordance with the 1964 Helsinki Declaration and its later amendments. The data used in this work were derived from the publicly available “Bilingualism and the Brain” dataset (OpenNeuro accession number: ds001796). The original data collection protocol was approved by the Research Ethics Committee of the University of Reading, and all participants provided written informed consent before study enrollment. Secondary analysis of this de-identified public dataset complies with ethical guidelines for human subjects’ research and open science data sharing policies.

We analyzed data from the Bilingualism and the Brain Dataset, which offers detailed language immersion variables integrated with multimodal neuroimaging assessments. Our sample comprised participants who acquired English as their second language and demonstrated high L2 proficiency, ensuring uniformity in linguistic background across the study cohort. Controlling proficiency levels allowed us to isolate the effects of immersion experience from language competence itself, thereby focusing specifically on how duration and quality of L2 environmental exposure influence glymphatic system function independent of skill-related variance.

### MRI Data Acquisition

Neuroimaging data were acquired on a 3T Siemens MAGNETOM Prisma_fit scanner equipped with a 32-channel head coil. The imaging protocol comprised three sequences: resting-state functional MRI (300 volumes; TR = 1,500 ms, TE = 30 ms; 2.1 × 2.1 × 2.0 mm voxel resolution; eyes-open condition), high-resolution T1-weighted MPRAGE structural imaging (0.7 mm slice thickness; TR = 2,400 ms, TE = 2.41 ms), and diffusion-weighted imaging (64 gradient directions; TR = 1,800 ms, TE = 70 ms; 2 mm isotropic voxels). All neuroimaging data are publicly available^126^.

### Language variables

Bilingual experience variables were derived from the Language and Social Background Questionnaire (LSBQ) ^127^. All aforementioned variables were log-transformed due to non-normal distributions and the expectation of non-linear adaptation patterns over time. L2 Age of Acquisition (L2 AOA) was obtained through self-report questionnaires, indicating the age at which participants began learning English. L2 Immersion duration was operationalized as the total time spent in the target language environment, calculated in months from participants’ arrival in the United Kingdom to the date of neuroimaging assessment. L2 Immersion Active Use (L2 Imm_Active) quantified the duration of active English engagement within the immersive environment, computed by first determining participants’ English usage percentage across four language modalities – reading, writing, speaking, and listening – then multiplying this percentage by total immersion months to yield an intensity-weighted exposure metric that distinguishes meaningful linguistic engagement from passive environmental presence^98^. For example, a participant residing in the UK for 100 months with 60% English usage would have L2 Imm_Active = 60 months.

L2 Immersion Passive Use (L2 Imm_Passive) represented the duration spent in the immersive environment with limited active English engagement, calculated by subtracting L2 Imm_Active from total immersion duration. This variable captured time potentially characterized by greater native language reliance or passive exposure rather than active communication. Additionally, we derived L2 Non-Immersion Active Use (L2 NonImm_Active) to quantify active English engagement outside immersive contexts. This measure was computed by first calculating participants’ lifelong L2 active use – determined by averaging English usage percentages across developmental stages from initial acquisition through testing – then subtracting L2 Immersion Active Use from this cumulative total. The resulting variable isolated English usage duration in non-immersive settings such as class.

*Note. All imaging data are displayed using sample subject 204 for demonstration purposes. CSF ‘production factory’ assessment: ChP ratio*

We segmented the choroid plexus for each participant using structural MRI^128^ (Figure 2.A). Specifically, T1-weighted images were registered to ICBM-MNI 152 standard space using ANTs software to reduce morphological variability. Choroid plexus segmentation was performed using a deep learning approach that demonstrated superior performance (Dice coefficient: 0.72, Hausdorff distance: 1.97mm) compared to atlas-based methods^129^. The model was trained on 64×64×64 voxel patches with data augmentation, and segmentation results were transformed back to native space for volume extraction. Raters independently verified segmentation quality across all participants, detailed described previously^138^. Total Intracranial Volume (TIV) was calculated using SPM12^130^ by summing gray matter, white matter, and CSF volumes, selected for its demonstrated consistency and low variability. Normalized choroid plexus volume was expressed as the percentage ratio of absolute choroid plexus volume to TIV.

**Figure 2.**
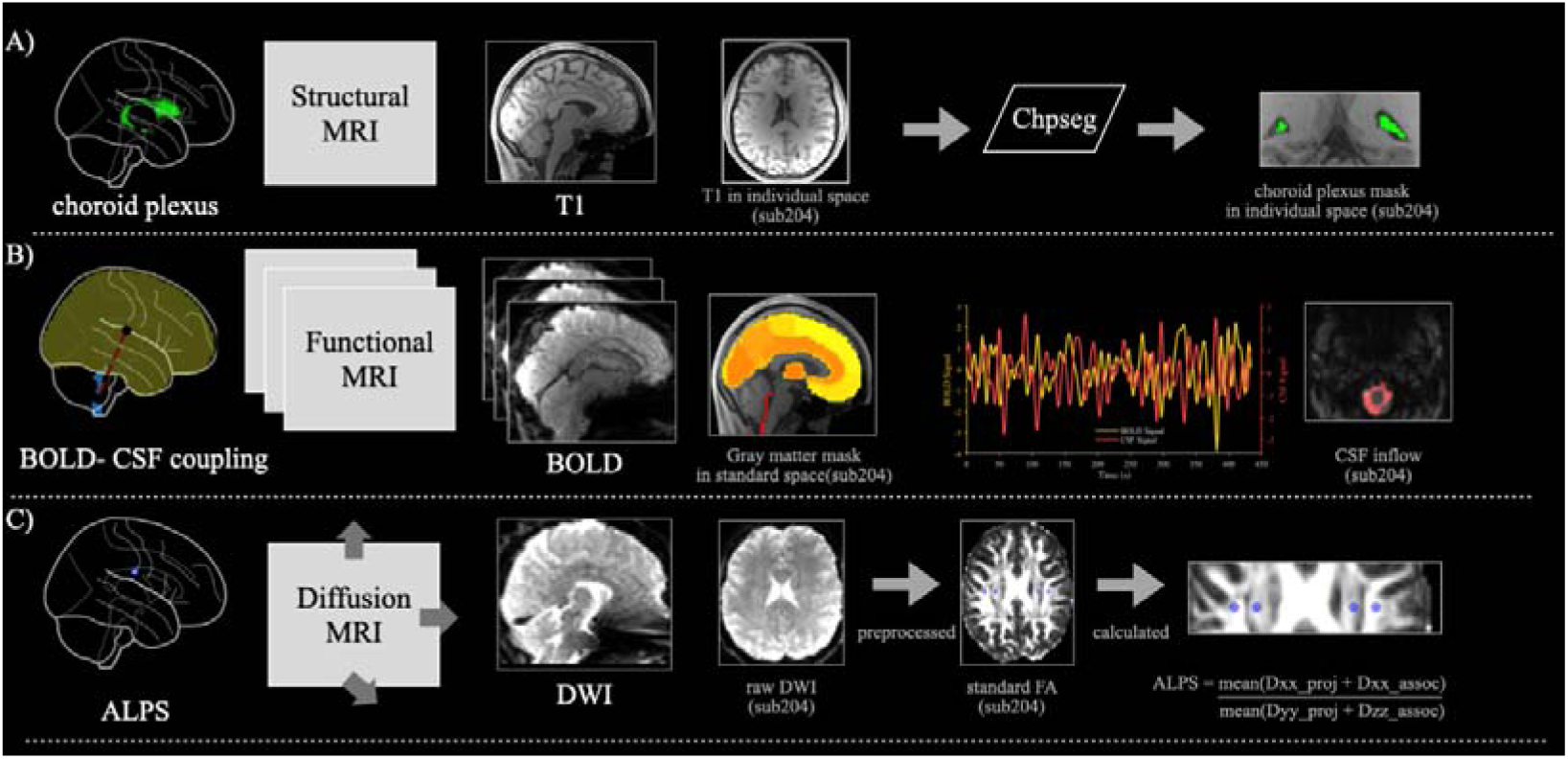
Protocol of multiple glymphatic measures. **A)** Choroid plexus segmentation via structural MRI. T1-weighted images in individual space (native) were processed through ChpSeg pipeline to generate choroid plexus masks in individual space (native). **B)** BOLD-CSF coupling via functional MRI. BOLD signals were extracted from gray matter mask and cross-correlated with CSF inflow signals to quantify coupling strength. **C)** ALPS index via diffusion MRI. Raw DWI images (native) were preprocessed, and standard FA images (native) were used to calculate the ALPS index through the formula: ALPS = mean(Dxx_proj + Dyy_proj) / mean(Dzz_assoc).

### CSF dynamic system assessment: BOLD-CSF coupling

As shown in Figure 2.B we extracted whole-brain gray matter BOLD time series and CSF signals from functional MRI data for each participant after preprocessing. Both signals underwent preprocessing with DPABI software^131^, including removal of the first 10 volumes, slice timing correction, motion correction, quadratic detrending, and bandpass filtering (0.01-0.1 Hz). Detailed preprocessing procedures have been reported previously^27,132^. For CSF signal extraction^18,133^, T1 images were registered to standard space via SPM12 to create CSF masks, with signals extracted from the first valid CSF mask slice in inferior-to-superior order. Each mask was validated against individual T1 images to ensure segmentation accuracy^27,132^. We conducted visual assessment of CSF mask segmentation quality for each participant, excluding participants whose segmentation failed due to incomplete cerebellar coverage in the field of view.

For whole-brain gray matter BOLD signal processing^18,133^, followed by spatial normalization to MNI space using DARTEL and smoothing with a 6mm FWHM Gaussian kernel. Final BOLD signals were extracted from gray matter regions defined by the Brainnetome Atlas (BNA246)^134^.

BOLD-CSF coupling was quantified using cross-correlation functions between whole-brain gray matter BOLD signals and CSF signals across time lags (−20s to 20s). The correlation strength at the negative peak was used as the coupling measure for each participant.

Cross-correlation between the negative temporal derivative of the global BOLD signal and CSF signal confirmed the expected inverse relationship^25^.

*Note: BOLD-CSF coupling strength is inversely related to coupling values—lower coupling values indicate stronger coordination between brain activity and CSF flow, reflecting more efficient glymphatic function*.

### Waste pathway permeability assessment: DTI-ALPS

As shown in Figure 2.C we calculated ALPS for each participant using diffusion MRI data processed entirely with FSL tools^135^. Raw diffusion data underwent preprocessing including brain extraction using BET, motion and eddy current correction via FSL’s “eddy_cuda” command. DTI fitting was performed using dti-fit on eddy corrected data to generate diffusivity parameter maps (FA, MD, AD, RD).

All DTI maps were registered to the JHU-ICBM-FA-1mm template using FLIRT for linear registration followed by FNIRT for nonlinear alignment. The outputs of FNIRT registration were manually inspected for each participant to ensure anatomical accuracy and to rule out potential image warping or distortions prior to ROI masking. Four regions of interest were defined from the JHU white matter atlas: bilateral superior corona radiata as projection fibers (proj) at coordinates L_SCR (53,47,40) and R_SCR (25,47,40), and bilateral superior longitudinal fasciculus as association fibers (assoc) at L_SLF (59,47,40) and R_SLF (19,47,40).

Directional diffusivity values (Dxx, Dyy, Dzz) were extracted from these ROIs for ALPS calculation. The ALPS index was computed as the bilateral mean ratio^136^:

ALPS = mean(Dxxproj + Dxxassoc))/(mean(Dyyproj + Dzzassoc)

*Note.where Dxxproj/assoc represents x-axis diffusivity in projection/association fibers, Dyyproj represents y-axis diffusivity in projection fibers, and Dzzassoc represents z-axis diffusivity in association fibers*.

### Data analysis

#### H1: Effects of L2 Immersion on Glymphatic Function

To examine associations between L2 immersion and glymphatic system functioning, we conducted partial correlation analyses between immersion duration and three glymphatic markers: BOLD-CSF coupling strength, choroid plexus ratio, and ALPS index, controlling for age, education level, and sex.

We tested a parallel mediation model examining whether L2 immersion duration influences ALPS index through two glymphatic markers: choroid plexus ratio and BOLD-CSF coupling strength. This model builds on previous findings establishing choroid plexus-to-ALPS pathways^137,138^ while exploring the understudied BOLD-CSF coupling-to-ALPS pathway. Given the glymphatic system’s integrated function and L2 immersion’s holistic brain effects, we modeled both pathways simultaneously:

***L2 immersion → {Choroid plexus ratio, BOLD-CSF coupling} → ALPS index***

We confirmed the two mediators were uncorrelated, supporting independent pathways. Path coefficients were estimated using ordinary least squares regression, controlling for age, education, and sex. Indirect effects were tested using bias-corrected bootstrap confidence intervals (n = 5000 samples, α = 0.05), with effects considered significant if 95% confidence intervals excluded zero^137^.

#### H2: Interactive Effects of L2 Immersion and L2 AOA on Glymphatic Function

To examine whether L2 AOA moderates the relationship between L2 immersion and glymphatic function, we conducted three interaction analyses. Three models examined distinct glymphatic outcomes: BOLD-CSF coupling (Model 1), choroid plexus ratio (Model 2), and ALPS index (Model 3). All models controlled for education, age, and sex.; and to ensure stable interaction estimates, we excluded data in unbalanced region^139^. Significant interactions were probed using simple slopes analysis^140^, with the Johnson-Neyman technique identifying critical AOA values where immersion effects became significant^141^.

#### H3: Active Immersion Specificity benefits glymphatic function

To test Hypothesis 3, we examined the differential effects of three L2 use patterns on glymphatic function: active L2 use within immersive environments, passive exposure within immersive environments, and active L2 use in non-immersive contexts. For each glymphatic marker—BOLD-CSF coupling strength, choroid plexus ratio, and ALPS index—we tested the association with each L2 use pattern separately, controlling for education, age, and sex. This approach allowed systematic comparison of how each L2 use pattern predicts glymphatic outcomes, thereby isolating the unique contribution of meaningful linguistic engagement in naturalistic environments:

***Glymphatic index = β□ + β□×L2_Variable + covariates + ε***

*Note:* where L2_Variable represents each of the three L2 use patterns examined across all glymphatic markers.

## Results

### Participants

As detailed in Table 1, the sample comprised 50 participants (39 females, 78%) with a mean age of 32.6 years (SD = 7.6). All participants demonstrated high L2 English proficiency (M = 9.4, SD = 1.0, range: 7.5–10.0 on a 10-point scale), ensuring that observed effects reflect immersion experience rather than language skill variability.

### BOLD-CSF coupling in high-proficiency L2 participants

BOLD-CSF coupling values were calculated at a 1.5-second lag with BOLD leading CSF signal (Figure 3.B). The average cross-correlation showed negative peaks at 1.5-3 second lags, while the first negative derivative exhibited significant peaks at zero lag. To examine coupling consistency across different L2 characteristics, we performed extreme group analyses (top and bottom 25%) for both L2AOA and L2immersion, confirming robust coupling patterns in early vs. late L2AOA groups and low vs. high L2immersion groups.

**Figure 3.**
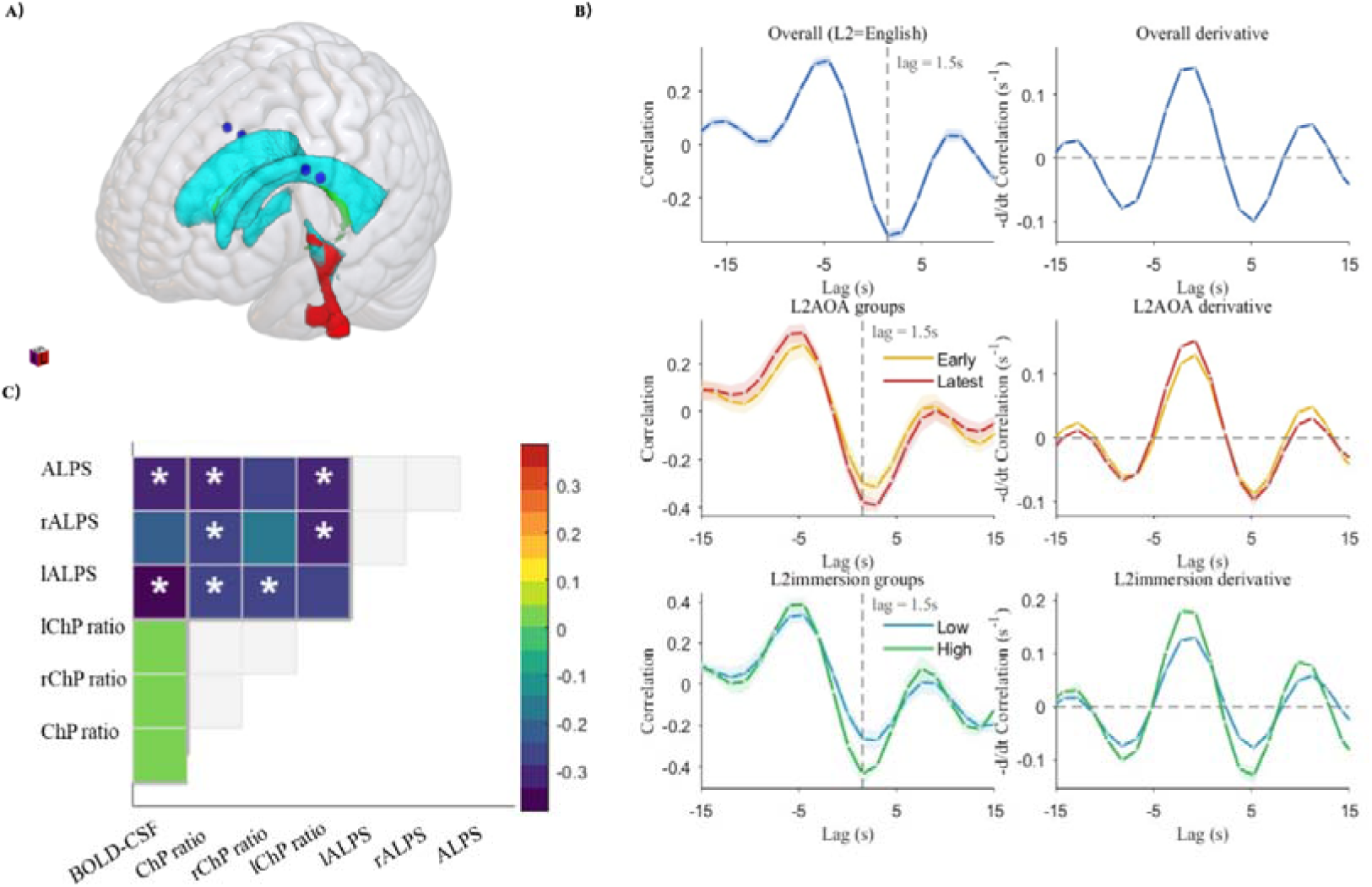
Glymphatic system function assessment. **A)** Three-dimensional schematic of glymphatic indices within the brain’s waste clearance network. **B)** Cross-correlation analysis between BOLD fluctuations and CSF dynamics, showing temporal coupling between neural activity and fluid movement across all L2-English participants and extreme groups (L2AOA: early vs. latest 25%; L2immersion: low vs. high 25%). **C)** Partial correlations among three glymphatic markers—BOLD-CSF coupling, choroid plexus ratio, and ALPS index—controlling for age, education, and sex.

### Relationships between glymphatic system indices in high-proficiency L2 participants

Partial correlations among glymphatic measures, controlling for education, age, and sex, are presented in Figure 3.C (detailed statistics in Table S1). Both BOLD-CSF coupling strength (r = −0.335, p < .05) and choroid plexus ratio (r = −0.325, p < .05) negatively correlated with composite ALPS, indicating that stronger brain-CSF coordination and reduced choroid plexus enlargement associate with enhanced perivascular flow efficiency.

Critically, BOLD-CSF coupling and choroid plexus ratio were uncorrelated (r = −0.029, p > .05), confirming independent pathways and supporting our parallel mediation model approach.

### Associations between L2 Immersion and Glymphatic Function (H1)

Our partial correlation analysis revealed that longer L2 immersion was associated with increased BOLD-CSF coupling strength (r = −0.32, p < 0.05) and shorter L2 immersion duration was associated with enlarged choroid plexus ratio (r = −0.39, p < 0.05). However, L2 immersion duration showed no significant direct correlation with ALPS after controlling for age, sex, and education level.

Despite the absence of direct associations with ALPS, parallel mediation model revealed that choroid plexus CSF ratio and BOLD-CSF coupling significantly mediated the relationship between L2 immersion and ALPS index (Figure 4.D). The indirect effects were 0.149 through choroid plexus ratio and 0.124 through BOLD-CSF coupling, with a total indirect effect of 0.273 (95% CI = 0.102-0.533).

**Figure 4.**
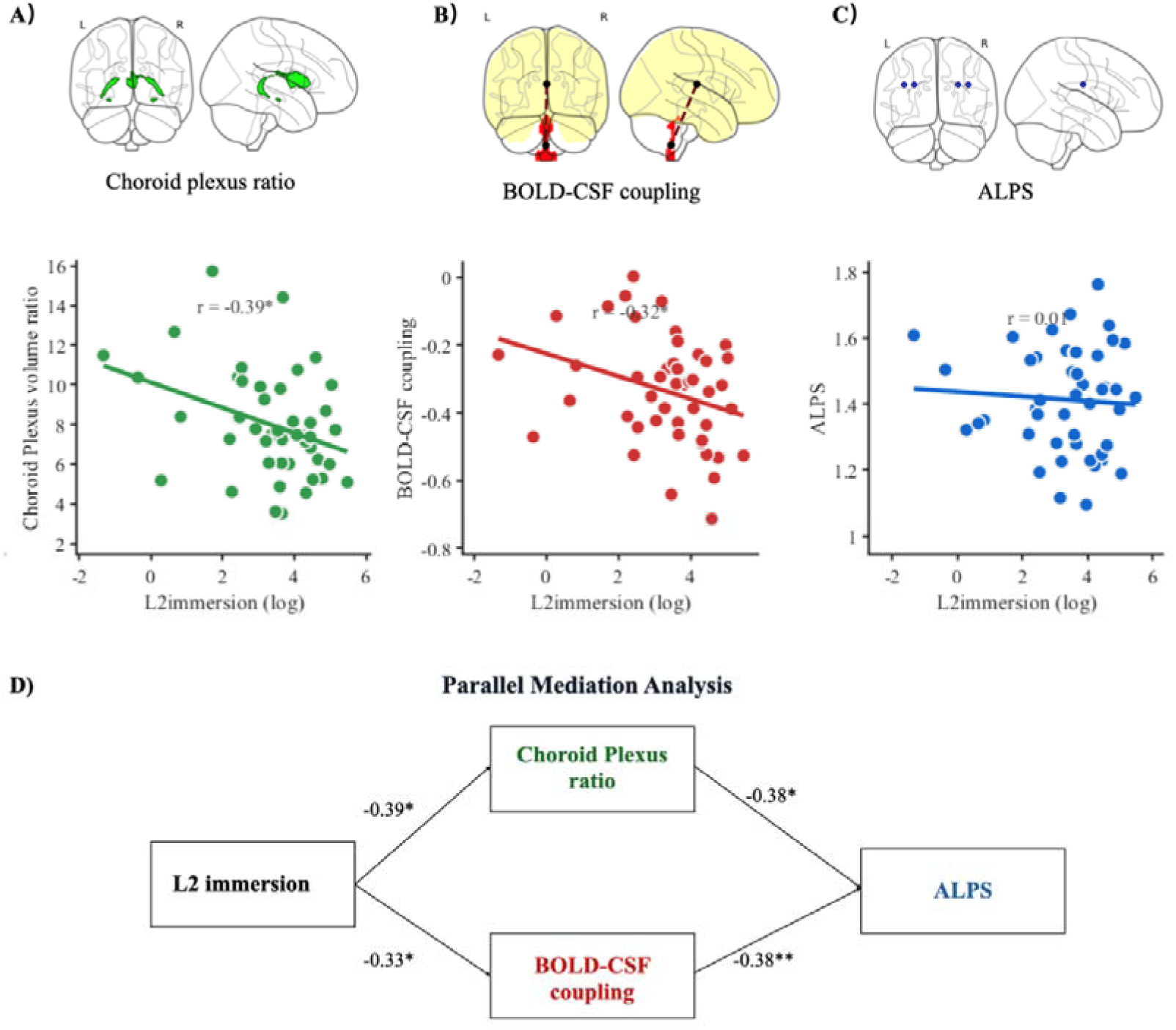
The relationship between L2 immersion and glymphatic function. **A)** L2 immersion has negative correlation with ChP enlargement **B)** L2 immersion has positive correlation with BOLD-CSF coupling intensity **C)** L2 immersion has no direct significant correlation with left ALPS. **D)** The mediation model of L2 immersion to left ALPS

Path coefficients showed that L2 immersion was negatively associated with choroid plexus CSF ratio (β = −0.392, p = 0.011) and BOLD-CSF coupling (standardized β = −0.328, p = 0.039). Both mediators were negatively associated with ALPS index (choroid plexus ratio: β = −0.380, p = 0.011; BOLD-CSF coupling: β = −0.379, p = 0.007).

### L2 Immersion × AOA interaction on brain-CSF coordination (H2)

As shown in Figure 5, L2 AOA significantly moderated the association between immersion duration and BOLD-CSF coupling strength (ΔR² = 0.189, F(1,33) = 10.75, p = 0.002; partial η² = 0.246). Simple slopes analysis in Figure 5.C showed that at early AOA (6.11 years), L2 immersion had no significant effect on BOLD-CSF coupling (B = −0.005, p = 0.879). At mean AOA (8.85 years), the effect approached significance (B = −0.044, p = 0.091), while at late AOA (12.94 years), L2 immersion significantly predicted enhanced BOLD-CSF coupling (B = −0.083, p = 0.014, 95% CI [−0.148, −0.018]). Johnson-Neyman analysis (Figure 5.C) identified a critical threshold at approximately 9.5 years of age (2.255 on the log-transformed scale), above which immersion duration significantly influenced BOLD-CSF coupling. In contrast, L2 AOA showed no significant moderating effects on the relationships between immersion duration and either choroid plexus ratios or ALPS index.

**Figure 5.**
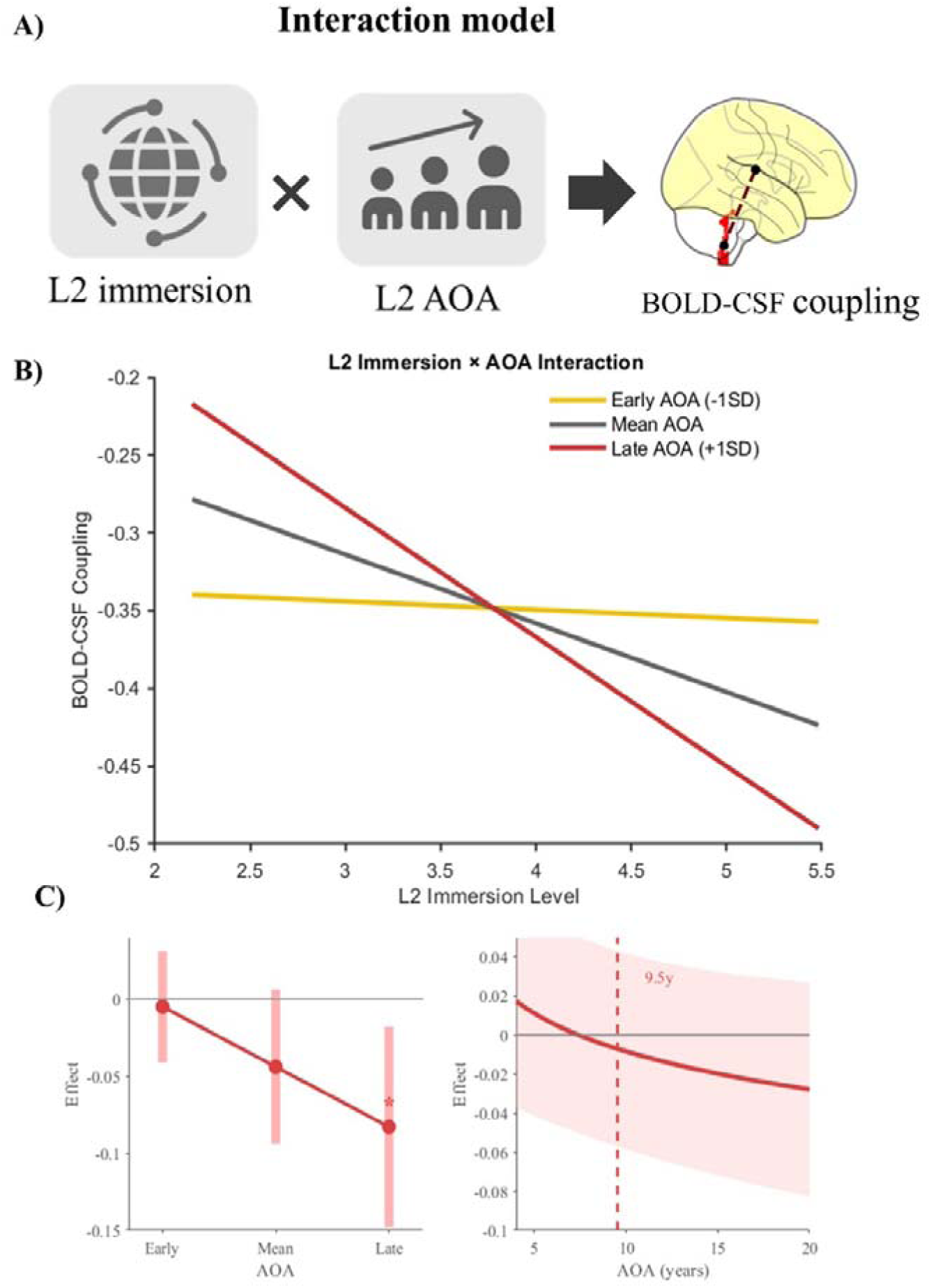
L2 immersion × AOA interaction effects BOLD-CSF coupling. **A)** Conceptual interaction model. **B)** Interaction plot showing immersion effects across early AOA (−1SD, yellow), mean AOA (gray), and late AOA (+1SD, red). **C)** Simple slopes analysis (left) and Johnson-Neyman significance regions (right) with critical threshold at 9.5 years (dashed line). *p < 0.05.

### Specific Effects of Active L2 Use in Immersion Environment (H3)

As shown in Figure 6, only L2_Imm_Active significantly predicted glymphatic function. Active immersive L2 use was negatively associated with BOLD-CSF coupling (β = −0.316, *p* = 0.047) and choroid plexus ratio (β = −0.351, *p* = 0.020), while L2_Imm_Passive and L2_NonImm_Active showed no significant associations across any glymphatic measures. The ALPS index was not significantly associated with any L2 use predictor.

**Figure 6.**
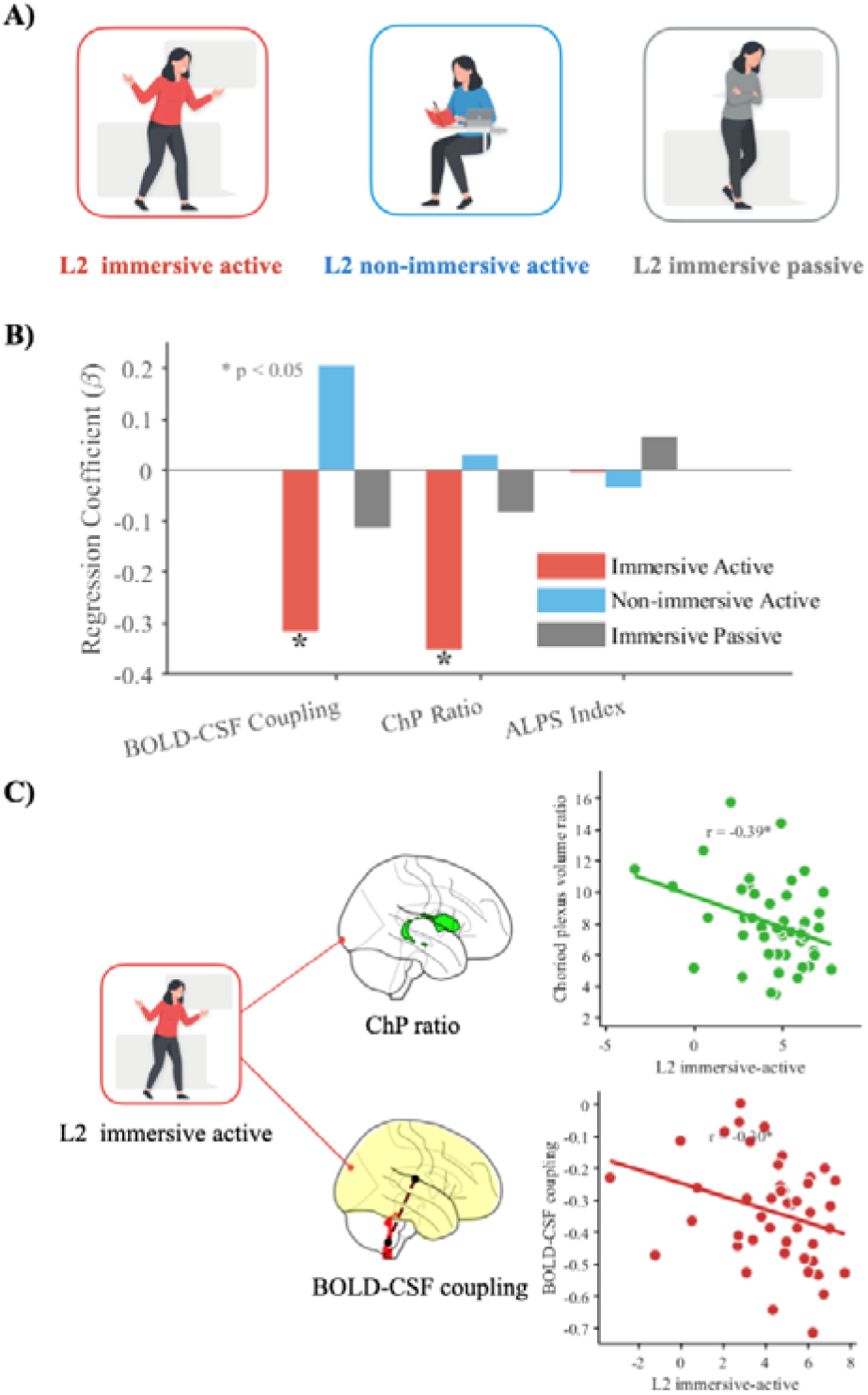
Specific effects of active L2 use in immersive environments on glymphatic function. **A)** Three L2 use patterns examined: immersive active, non-immersive active, and immersive passive. **B)** Standardized regression coefficients (β) for associations between L2 use patterns and glymphatic measures. Only L2 immersive active (red bars) showed significant negative associations with BOLD-CSF coupling and choroid plexus (ChP) ratio (*p < 0.05). **C)** Scatter plots illustrate significant relationships between L2 immersive active use and glymphatic markers, showing reduced ChP ratio and increased BOLD-CSF coupling strength.

## Discussion

Our study provides the first empirical evidence linking second language immersion experience with better glymphatic system function in healthy young adults. Our findings indicate that L2 immersion duration may be associated with optimized glymphatic function across multiple physiological pathways, including improved brain-CSF coordination, reduced choroid plexus enlargement, and enhanced perivascular flow dynamics. These results extend our understanding of bilingualism’s neuroprotective effects beyond traditional cognitive and structural neuroimaging outcomes to potentially encompass fundamental physiological processes that maintain brain health.

### Validity of Multimodal Glymphatic Assessment

As glymphatic function represents an integrated system, individual biomarkers face interpretation challenges despite widespread validation^35^. ALPS measures radial water movement at the lateral ventricular level and cannot independently reflect overall glymphatic function, prompting calls for multi-method approaches^19^. Without a gold standard, this study combined three modalities for glymphatic assessment, revealing consistent associations in healthy populations (Figure 3.C). Higher ALPS correlated with smaller choroid plexus volume and stronger BOLD-CSF coupling. These directional associations align with underlying biological hypotheses^137,138^, supporting comprehensive interpretation of our findings. Therefore, these convergent findings may warrant discussion in terms of their underlying biological significance: improved brain-CSF coordination, reduced choroid plexus enlargement, and enhanced perivascular flow dynamics.

### L2 Immersion may Enhance BOLD-CSF coupling as a Beneficial Adaptation

Partial correlation analyses revealed that longer L2 immersion duration significantly associated with higher BOLD-CSF coupling strength. This relationship may reflect beneficial adaptations in bilinguals. First, our previous work established BOLD-CSF coupling as a valid glymphatic biomarker^27,132^, with neurodegenerative patients showing reduced coupling strength and associated protein deposition^17,27^. Building on this foundation, stronger coupling in healthy populations appears to represent adaptive reserve capacity rather than merely normal function. This is evidenced by enhanced coupling in young adults following sleep deprivation (absent in older adults)^142^ and stronger coupling in younger individuals with better viral infection recovery outcomes. Similarly, L2 immersion represents a substantial challenge, and individuals achieving fluency may share enhanced BOLD-CSF coupling as a reserve mechanism with young adults who successfully adapt to sleep deprivation or recover from infections.

### L2 Immersion May Optimized Choroid Plexus Structure as an Aging Resilience

This study revealed that shorter L2 immersion duration significantly associated with larger choroid plexus ratios (Figure 4.A), suggesting that L2 immersion may decelerate choroid plexus aging processes. The directional relationship between choroid plexus morphology and L2 immersion aligns with our hypothesis, as choroid plexus enlargement likely reflects inflammatory swelling^143^ consistently reported across various brain disorders, establishing it as a robust pathological biomarker^26^. Moreover, choroid plexus volume correlates with aging trajectory and serves as a potential aging biomarker^13,14^. Studies show physical exercise prevents neurovascular degeneration through choroid plexus modulation^145^, while bilingualism similarly represents continuous cognitive exercise within the brain. Observational studies demonstrate bilingualism experience confers aging resistance^146^. These findings suggest choroid plexus may quantify the brain anti-aging benefits of foreign language exposure in the future.

### L2 Immersion Indirect Effects on ALPS

Our mediation analysis revealed that L2 immersion may influence ALPS indirectly through choroid plexus ratio and BOLD-CSF coupling, with significant total indirect effects but no direct pathway. The negative association between choroid plexus and ALPS aligns with previous clinical studies, as choroid plexus enlargement may impair subsequent CSF-ISF exchange capacity^137,138^. While BOLD-CSF coupling and ALPS associations have been reported^147^, this study first examined potential directionality through mediation analysis, possibly reflecting stronger CSF inflow enhancing perivascular clearance efficiency. These biological relationships lie beyond the scope of this study and require validation through larger causal studies.

The absence of direct L2 immersion-ALPS associations may reflect that ALPS is particularly sensitive to severe pathology and aging. ALPS consistently decreases in neurodegenerative diseases^148–150^ and serves as a brain reserve index in elderly populations^151^. Given our young healthy bilingual sample, ALPS changes may not yet be detectable, suggesting future research should examine ALPS-brain reserve relationships in older bilingual cohorts to establish clearer associations. The downstream positioning of ALPS in our mediation model may reflect both inherent glymphatic physiological mechanisms and the sequential nature of language experience effects, potentially paralleling the ordered dynamics observed in cortical reorganization^152^.

### Age of Acquisition Moderates Brain-CSF Coordination Effects

The moderation analysis revealed that L2 AOA significantly influenced the relationship between immersion duration and BOLD-CSF coupling strength. The AOA-dependent effects on BOLD-CSF coupling strength point to age-sensitive mechanisms, with late learners demonstrating stronger associations than early learners. Individuals who began acquiring English after approximately age 9.5 showed significant immersion-related improvements in brain-CSF coordination, whereas earlier learners exhibited no such relationship. This pattern suggests that late bilinguals may require or engage different neurophysiological mechanisms to achieve functional outcomes in immersive environments, potentially recruiting compensatory adaptations that include enhanced glymphatic efficiency.

The absence of AOA moderation effects on choroid plexus morphology indicates that structural optimization of CSF production sites occurs independently of acquisition timing, suggesting that these adaptations represent universal responses to immersive experience regardless of when language learning began. In contrast, the AOA-dependent effects on brain-CSF coupling dynamics point to age-sensitive mechanisms that differ between early and late bilingual acquisition. Late learners may experience greater neurovascular plasticity in response to immersion demands, as their mature brain systems adapt to novel linguistic challenges through enhanced coordination between neural activity and fluid circulation.

These findings partially align with critical period frameworks while revealing unexpected complexity^153^. This pattern may initially appear counterintuitive but considering that our sample’s mean L2 subjective proficiency was 9.4/10, participants demonstrated consistently high fluency levels. This may have two explanatory reasons: 1) later learners who achieve the same level of fluency require greater effort, or 2) later learners face greater challenges when encountering overseas L2 immersion environments at later stages. For both cases, we postulate that the greater effort later learners put into place induces optimalized CSF coupling dynamics.

The first explanation aligns with the highlight the complexity of L2 AOA effects^118^, suggesting that age acts as a proxy for interacting, suggesting that age acts as a proxy for interacting cognitive and social constraints rather than a simple maturational cutoff cognitive and social constraints rather than a simple maturational cutoff. According to this framework, later learners who achieved equivalent proficiency levels may recruit additional cognitive resources and employ distinct processing strategies may drive more pronounced neurovascular coupling changes that manifest as stronger brain-CSF coordination improvements. Alternatively, these patterns can be interpreted through the Dynamic Restructuring Model (DRM), which posits that brain adaptations to bilingualism are experience dependent. This interpretation is supported by our previous research showing that virus-infected patients exhibited enhanced BOLD-CSF coupling strength as a stress-responsive compensatory mechanism, with greater compensation associated with better recovery outcomes. Similarly, the enhanced glymphatic sensitivity observed in late learners may represent adaptive physiological responses to immersion-related stress that ultimately support successful language functioning in challenging environments. Alternatively, early bilinguals may have already optimized these systems during initial acquisition, leaving less room for immersion-related enhancement.

Rather than simply showing diminished plasticity, late learners demonstrate enhanced sensitivity to immersion duration in specific glymphatic domains. The enhanced BOLD-CSF coupling observed in our sample may represent a proactive physiological adaptation to sustained cognitive load. The intense demands of active bilingual immersion may trigger early-stage optimization of waste clearance mechanisms. This suggests that bilingual experience functions as a potent physiological driver that strengthens brain maintenance systems before the onset of age-related decline.“

### Active Engagement Within Immersive Contexts Drives Better Glymphatic Function

Active immersive engagement significantly predicted both enhanced BOLD-CSF coupling and reduced choroid plexus ratios, indicating coordinated functional and structural optimization of CSF dynamics^12,28,29,142^. In contrast, passive exposure and non-immersive active use showed no such associations, underscoring the critical importance of combining ecological linguistic demands with meaningful communicative participation. That is consistent with the *Interaction Hypothesis* in linguistics, which emphasizes that authentic communicative practice, rather than passive exposure, is essential for sustained language learning and cognitive progress^156^.

The unique effects on both glymphatic measures suggest that genuine linguistic engagement within naturalistic environments triggers inflammatory modulation, vascular remodeling, or metabolic changes that optimize CSF dynamics. Passive immersion lacks the communicative pressure necessary for these adaptations, while non-immersive active use may lack the ecological validity and multisensory integration of natural communication settings. ALPS indices showed limited sensitivity as downstream effects in young healthy populations, potentially reflecting statistical power constraints and responsiveness to unmeasured bilingual dimensions like code-switching frequency or linguistic distance.

### Bilingual Glymphatic Advantages as Neuroplasticity Extension

While cognitive and brain advantages of bilingualism are well-documented^10,11,14,30,151^, our findings suggest neuroplasticity benefits extend beyond neuronal structures to fundamental brain systems like the glymphatic system.

The sustained cognitive demands of bilingual environments may trigger adaptations that optimize CSF dynamics. Enhanced BOLD-CSF coupling represents adaptive compensation when the brain faces increased waste removal demands^142^. L2 immersion elevates neuronal metabolic activity through heightened cognitive demands, generating increased waste products^157^ that potentially trigger compensatory glymphatic enhancement. Molecular studies demonstrate that exercise-derived metabolites directly target choroid plexus epithelial cell metabolic enzymes, providing neuroprotection against aging^145^. Bilingual cognitive processing, similar to exercise, requires sustained high energy consumption through language monitoring, executive control, and code-switching, potentially triggering comparable adaptive metabolic restructuring^158,159^. Our study identified L2 immersion glymphatic advantages through three complementary measures. Given glymphatic dysfunction’s role in neurodegeneration and aging, understanding bilingual advantages may inform approaches to combat disease progression. Future mechanistic studies are needed.

### Mechanistic Implications and Neurobiological Pathways

While cognitive advantages of bilingualism are well-documented^49–51^, our findings reveal that neuroplasticity benefits extend beyond neuronal structures to fundamental brain clearance systems. L2 immersion demonstrates glymphatic advantages through three complementary measures: enhanced BOLD-CSF coupling, reduced choroid plexus ratios, and improved perivascular flow dynamics.

These effects likely arise through three interconnected mechanisms. Increased waste removal demands drive enhanced BOLD-CSF coupling as adaptive compensation^142^ when L2 immersion elevates neuronal metabolic activity through heightened cognitive demands, generating increased waste products^157^ that potentially trigger compensatory glymphatic enhancement. Enhanced metabolic capacity emerges through sustained bilingual experience triggering metabolic adaptations similar to exercise^145^, targeting choroid plexus epithelial cell enzymes and anti-inflammatory pathways to maintain efficient CSF production and clearance capacity. Optimized neurovascular coordination occurs through strengthened subcortical nuclei and enhanced cholinergic function that better orchestrate neurovascular coupling^160^, promoting synchronized brain-CSF dynamics via improved slow-wave activity regulation and widespread neural network remodeling.

### Implications for Neuroprotection and Cognitive Reserve

Despite the fact that our study focused on healthy adults, our findings may also provide novel mechanistic insights into how bilingual experience may potentially confer protection against cognitive decline and neurodegeneration. Previous research has documented delayed dementia onset in bilinguals and preserved cognitive function despite equivalent neuropathological burden, suggesting cognitive reserve mechanisms that compensate for disease-related damage^161^. However, the physiological pathways underlying these protective effects have remained unclear. Our results indicate that enhanced glymphatic function may represent a critical biological mechanism through which bilingual experience builds brain resilience.

Impaired glymphatic clearance contributes fundamentally to neurodegenerative disease progression by allowing toxic protein accumulation, including amyloid-beta and tau aggregates characteristic of Alzheimer’s disease^119–125^. If bilingual immersion experience enhances waste removal efficiency during midlife, as our results suggest, this could reduce pathological protein burden over decades, delaying clinical symptom emergence even as underlying disease processes advance. The brain-CSF coordination improvements and choroid plexus optimization we observed in relatively young participants (mean age 32.6 years) may establish protective physiological patterns that compound across the lifespan, contributing to the cognitive reserve advantages documented in older bilingual populations.

The indirect pathway from immersion through glymphatic components to perivascular flow suggests that bilingual experience initiates cascading adaptations across integrated brain systems. Rather than producing isolated local changes, sustained linguistic engagement appears to promote system-wide optimization of fluid dynamics, metabolic efficiency, and inflammatory regulation. These coordinated adaptations may collectively enhance brain resilience to multiple insults, including not only protein aggregation but also vascular injury, inflammatory challenges, and metabolic stress that contribute to cognitive aging.

### Limitations and Future Directions

Several limitations warrant consideration. The cross-sectional design precludes causal inferences, requiring longitudinal studies to establish directionality and temporal dynamics. Sample size constraints limited statistical power, while the homogeneous sample of high-proficiency L2 English learners restricts generalizability across linguistic combinations and cultural contexts. Our glymphatic measures provide indirect assessments of waste clearance function; future studies incorporating contrast-enhanced MRI, CSF biomarkers, or direct fluid visualization methods would strengthen mechanistic understanding. The mediation models represent preliminary exploration rather than definitive causal evidence, and unexplored variables including sleep quality, physical activity, and stress exposure may confound observed relationships. Additionally, our assessment of active versus passive engagement relied on subjective self-report scales; future research should develop more objective methods to quantify actual language use. The specificity analysis revealed significant effects only for choroid plexus morphology, suggesting refined measurement approaches capturing code-switching density, communicative complexity, and social interaction quality may reveal more nuanced relationships. Lastly, our current analysis did not directly measure lifestyle factors such as sleep quality or physical activity, which are known modulators of glymphatic function. Future longitudinal studies incorporating objective sleep data across the lifespan will be essential to clarify how bilingual immersion interacts with these factors to foster long-term cognitive resilience.

## Conclusion

This study provides the first empirical evidence that L2 immersion experience associates with enhanced glymphatic function in healthy young adults. Longer immersion duration predicted improved brain-CSF coordination, optimized choroid plexus morphology, and enhanced perivascular flow, suggesting coordinated adaptations across the integrated waste clearance network. Age of acquisition moderated brain-CSF coupling effects, with late learners showing stronger improvements, while active immersive engagement specifically influenced choroid plexus and BOLD-CSF coupling optimization. These findings extend bilingualism’s neuroprotective effects beyond cognitive outcomes to fundamental physiological processes maintaining brain health. Enhanced glymphatic function may represent a critical mechanism through which sustained bilingual engagement builds cognitive reserve and delays neurodegenerative decline, providing novel insight into accessible approaches for promoting brain health. Future longitudinal research with diverse populations and comprehensive glymphatic assessment will be essential for establishing causality and translating findings into evidence-based interventions for cognitive aging prevention.

## Appendix

**Table 1.** Participants Demographic and Language Background Characteristics (N = 50)

| Variable | M | SD | Range |
| --- | --- | --- | --- |
| Sex | Female: 39 (78.0%); Male: 11 (22.0%) |  |  |
| Age | 32.6 | 7.6 | 22.0-52.0 |
| L2 Age of Acquisition | 9.2 | 3.5 | 4.0-20.0 |
| L2 Immersion (Months) | 58.3 | 55.9 | 0.3-240.3 |
| L2 Proficiency | 9.4 | 1.0 | 7.5-10.0 |
| L2 Imm_Active (Months) | 47.1 | 47.9 | 0.1-210 |
| L2 Imm_Passive (Months) | 11.2 | 11.9 | 0.001-45.1 |
| L2 NonImm_Active (Month) | 70.2 | 60.7 | 0.001-305 |
| L2 AOA (log) | 2.15 | 0.38 | 1.39-3 |
| L2 Immersion (log) | 4.37 | 1.51 | -1.34-5.48 |
| L2 Imm_Active (log) | 4.44 | 2.33 | -3.34-7.72 |
| L2 Imm_Passive (log) | 1.44 | 3.82 | -6.64-5.5 |
| L2 NonImm_Active (log) | 5.4 | 2.22 | -6.64-8.25 |

**Note.** M = Mean; SD = Standard Deviation.

**Table 2.** Participants Glymphatic Function Indexes.

| Variable | M | SD | Range | N |
| --- | --- | --- | --- | --- |
| BOLD-CSF coupling | -0.34 | 0.16 | -0.71-0.00 | 46 |
| ALPS | 1.41 | 0.16 | 1.09-1.76 | 50 |
| left ALPS | 1.4 | 0.17 | 1.11-1.81 | 50 |
| right ALPS | 1.41 | 0.17 | 1.03-1.72 | 50 |
| ChP ratio (%) | 1.49 | 0.49 | 0.6-2.52 | 50 |
| Left ChP ratio (%) | 0.7 | 0.25 | 0.27-1.28 | 50 |
| Right ChP ratio (%) | 0.79 | 0.26 | 0.33-1.34 | 50 |

**Note.** M = Mean; SD = Standard Deviation.

**Table S1.** Partial Correlations Among Glymphatic Function Measures.

| Variable | 1 | 2 | 3 | 4 | 5 | 6 | 7 |
| --- | --- | --- | --- | --- | --- | --- | --- |
| 1. BOLD-CSF coupling | – |  |  |  |  |  |  |
| 2. lALPS | -0.386* | – |  |  |  |  |  |
| 3. rALPS | -0.231 | 0.670*** | – |  |  |  |  |
| 4. ALPS | -0.335* | 0.915*** | 0.912*** | – |  |  |  |
| 5. ChP ratio | -0.029 | -0.304* | -0.289* | -0.325* | – |  |  |
| 6. rChP ratio | -0.015 | -0.294* | -0.2 | -0.271 | 0.944*** | – |  |
| 7. lChP ratio | -0.041 | -0.278 | -0.348* | -0.337* | 0.937*** | 0.711*** | – |

*Note.* Partial correlations controlling for education, age, and sex.
\*p < .05. \*\*p < .01. \*\*\*p < .001.

**Table S2.**
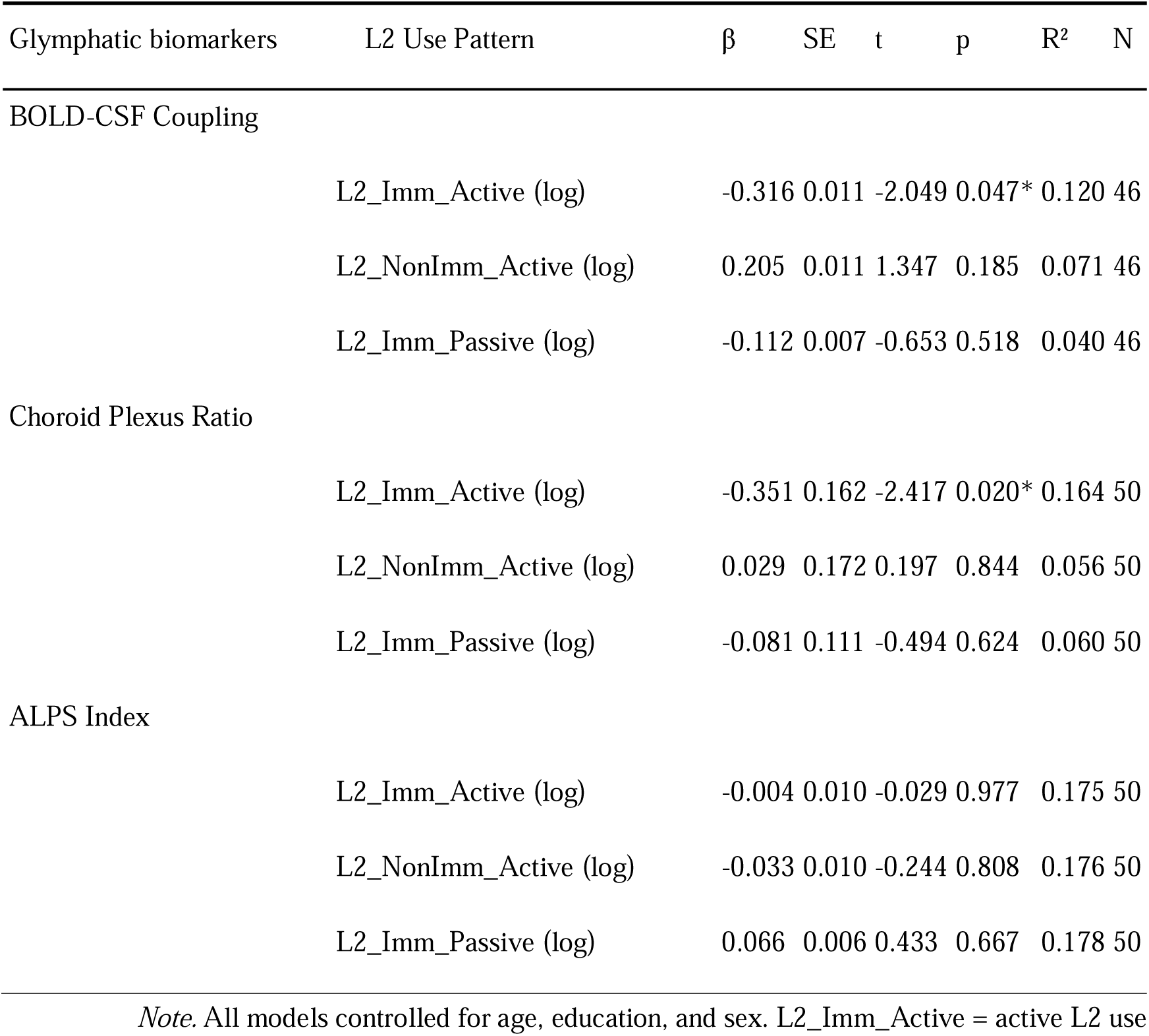
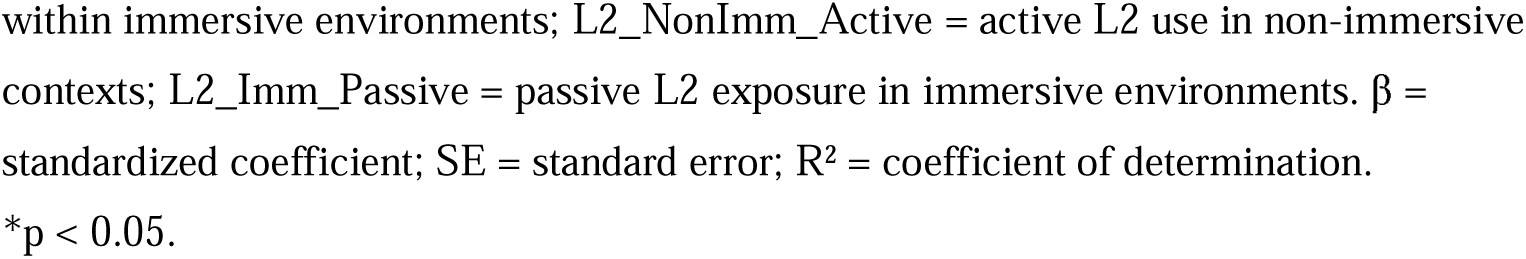
Associations Between L2 Use Patterns and Glymphatic Function Markers Glymphatic biomarkers L2 Use Pattern β SE t p R² N.

| Glymphatic biomarkers | L2 Use Pattern | $\beta$ | SE | t | p | R<sup>2</sup> | N |
| --- | --- | --- | --- | --- | --- | --- | --- |
| BOLD-CSF Coupling |  |  |  |  |  |  |  |
|  | L2_Imm_Active (log) | -0.316 | 0.011 | -2.049 | 0.047* | 0.120 | 46 |
|  | L2_NonImm_Active (log) | 0.205 | 0.011 | 1.347 | 0.185 | 0.071 | 46 |
|  | L2_Imm_Passive (log) | -0.112 | 0.007 | -0.653 | 0.518 | 0.040 | 46 |
| Choroid Plexus Ratio |  |  |  |  |  |  |  |
|  | L2_Imm_Active (log) | -0.351 | 0.162 | -2.417 | 0.020* | 0.164 | 50 |
|  | L2_NonImm_Active (log) | 0.029 | 0.172 | 0.197 | 0.844 | 0.056 | 50 |
|  | L2_Imm_Passive (log) | -0.081 | 0.111 | -0.494 | 0.624 | 0.060 | 50 |
| ALPS Index |  |  |  |  |  |  |  |
|  | L2_Imm_Active (log) | -0.004 | 0.010 | -0.029 | 0.977 | 0.175 | 50 |
|  | L2_NonImm_Active (log) | -0.033 | 0.010 | -0.244 | 0.808 | 0.176 | 50 |
|  | L2_Imm_Passive (log) | 0.066 | 0.006 | 0.433 | 0.667 | 0.178 | 50 |

*Note.* All models controlled for age, education, and sex. L2\_Imm\_Active = active L2 use
within immersive environments; L2\_NonImm\_Active = active L2 use in non-immersive contexts; L2\_Imm\_Passive = passive L2 exposure in immersive environments. $\beta$ = standardized coefficient; SE = standard error; $R^2$ = coefficient of determination. \* $p < 0.05$ .

## Data Availability

The data supporting this study findings are available from the OpenNeuro repository under the accession number ds001796 (https://doi.org/10.18112/OPENNEURO.DS001796.V1.7.0). Supplementary tables are included in the manuscript, and custom analysis code is available from the corresponding author on reasonable request.

https://openneuro.org/datasets/ds001796/versions/1.7.0

